# Pediatric Out-of-Hospital Cardiac Arrest Incidence During and After COVID-19 Social Distancing in Korea: A Nationwide 16-Year Interrupted Time-Series Analysis

**DOI:** 10.64898/2026.11.11.26367882

**Authors:** In Young Choi, Yonghyuk Jeon, In Kyung Lee, Kyunghoon Kim

## Abstract

**Background:** Pediatric out-of-hospital cardiac arrest (OHCA) is a rare but devastating event. However, the extent to which large-scale public health interventions, such as COVID-19 social distancing policies, affected the incidence of pediatric OHCA remains unclear.

**Methods:** We conducted a nationwide interrupted time-series analysis using data from the Korea Out-of-Hospital Cardiac Arrest Registry (2008–2023). The primary outcome was the monthly incidence of OHCA among individuals aged 0–18 years. The study period was divided into three phases: pre-intervention (January 2008–February 2020), social distancing (March 2020–April 2022), and post-lifting (May 2022–December 2023). We used segmented Poisson regression, adjusted for population size and seasonal variation, to estimate changes in monthly incidence at each policy change and reported the results as incidence rate ratios (IRRs). To determine whether these changes differed by age, we compared children with adults (19–64 years) and elderly individuals (≥65 years).

**Results:** Of the 456,839 registry entries, 10,965 were pediatric OHCAs, of which 6,891 underwent resuscitation in the emergency department. The crude incidence was 7.04, 7.14, and 8.35 per 100,000 person-years in the three phases, respectively. Before the pandemic, incidence rose slightly (annual IRR 1.006 [95% CI 1.001–1.012]; P = 0.032). At the implementation of social distancing, incidence fell immediately (IRR 0.926 [95% CI 0.865–0.991]; P = 0.027), without a significant change in slope (P = 0.081). After restrictions were lifted, incidence rose immediately (IRR 1.138 [95% CI 1.048–1.235]; P = 0.002) and then declined gradually (monthly IRR 0.993 [95% CI 0.988–0.999]; P = 0.023). These changes were confined to children aged 1–11 years (implementation IRR 0.812 [95% CI 0.666–0.991]; post-lifting IRR 1.562 [95% CI 1.278–1.909]). The pattern of change differed significantly by age group (P < 0.001), mainly because incidence rose in children, but not in adults or elderly individuals, after restrictions were lifted.

**Conclusions:** Nationwide social distancing was associated with a transient decrease in pediatric OHCA incidence, which was concentrated among children aged 1–11 years and was followed by a rebound above the expected level after restrictions were lifted. Surveillance of pediatric OHCA should account for abrupt changes in children’s daily environments, including the closure and reopening of schools and childcare facilities.

## Introduction

Pediatric out-of-hospital cardiac arrest (OHCA) is an uncommon but highly fatal condition with persistently poor outcomes (1–3). Unlike adult OHCA, pediatric cases are often related to non-cardiac etiologies, including respiratory failure, trauma, and congenital conditions (4, 5). Despite its clinical importance, the epidemiology of pediatric OHCA remains poorly characterized, largely because of its low incidence and clinical heterogeneity and the limited availability of large-scale datasets. Most existing studies have relied on cross-sectional or pre–post comparisons and lack a robust evaluation of temporal changes (6). Pediatric OHCA is driven substantially by external and potentially modifiable risk factors, including trauma, choking, and infection-related respiratory failure (4, 5, 7). These conditions are influenced by patterns of physical activity, environmental exposure, and access to healthcare (8–10). Population-level interventions that affect mobility, social interaction, and healthcare-seeking behavior may therefore influence the incidence of pediatric OHCA.

The COVID-19 pandemic prompted large-scale public health interventions, including nationwide social distancing policies, which substantially altered daily activities, mobility, and access to healthcare (8–10). These changes may have influenced the occurrence of pediatric OHCA through altered injury patterns, exposure to infectious diseases, and changes in healthcare-seeking behavior. However, prior studies have primarily compared the pre- and post-pandemic periods without isolating the policy phase itself or the period after the policies were withdrawn, which has limited the ability to assess the specific impact of social distancing measures (11).

Korea provides a suitable setting in which to evaluate the policy-level impact of social distancing on pediatric OHCA (12). From March 2020 to April 2022, the Korean government implemented a formal, tiered social distancing framework with clearly defined phases, and most restrictions were lifted on April 18, 2022. This structured policy timeline enables the precise delineation of intervention periods, in contrast to settings in which restrictions were applied inconsistently or varied across regions.

In addition, the Korea Out-of-Hospital Cardiac Arrest Registry (KOHCAR) is a government-mandated nationwide registry that has systematically captured all emergency medical services (EMS)-assessed OHCA cases since 2008 (13). By integrating prehospital and in-hospital data according to standardized Utstein definitions, KOHCAR provides a robust platform for the population-level analysis of rare events such as pediatric OHCA. Nonetheless, analyses of KOHCAR data focusing on pediatric patients remain limited.

In this study, we used data from a 16-year nationwide registry to evaluate long-term temporal trends in the incidence of pediatric OHCA, estimate the changes associated with both the implementation and the lifting of COVID-19 social distancing policies, and examine whether these changes differed across age groups.

## Methods

### Study design and data source

We conducted a nationwide, population-based interrupted time-series (ITS) study using KOHCAR, which is maintained by the Korea Disease Control and Prevention Agency (KDCA) in collaboration with the National Fire Agency (13). KOHCAR includes all EMS-assessed OHCA cases in Korea regardless of etiology and integrates prehospital EMS data with in-hospital data obtained through a standardized review of medical records based on the core Utstein variables (14). All data were de-identified before analysis and were accessed under an official data use agreement with the KDCA. This study is reported in accordance with the Strengthening the Reporting of Observational Studies in Epidemiology (STROBE) and REporting of studies Conducted using Observational Routinely-collected health Data (RECORD) statements (15).

### Study population

All registry entries from January 2008 to December 2023 were screened (N = 456,839). After the exclusion of 18 entries with unknown age, the remaining cases were classified as pediatric (0–18 years; n = 10,965), adult (19–64 years; n = 183,181), or elderly (≥65 years; n = 262,675) (Supplementary Figure 1). Two study populations were analyzed. The incidence population comprised all EMS-assessed OHCA cases in each age group, consistent with the Utstein recommendations for population-based incidence (14). The clinical cohort comprised cases in which cardiopulmonary resuscitation (CPR) was attempted in the emergency department (ED). For the pediatric clinical cohort, we excluded cases with an unrecorded ED CPR status (n = 719) and those in which CPR was not attempted in the ED (n = 3,355; dead on arrival, 3,275; do-not-attempt-resuscitation order, 76; transferred, 3; reason unknown, 1), leaving 6,891 patients for analysis. The adult (n = 103,195) and elderly (n = 129,366) clinical cohorts were defined using the same criteria.

### Exposure definition

To evaluate the impact of COVID-19 social distancing policies, we divided the study period into three predefined phases according to the timing of national policy implementation: Phase I (pre-intervention; January 2008–February 2020), Phase II (social distancing; March 2020–April 2022), and Phase III (post-lifting; May 2022–December 2023). Because restrictions were lifted in mid-April 2022, May 2022 was the first full calendar month thereafter and was therefore designated as the start of Phase III.

### Variables

The variables included demographic characteristics (age, sex, and comorbidities), arrest characteristics (location, etiology [disease-related, with subtype, or injury-related, with mechanism], intent, and first responder type), prehospital factors (bystander CPR and time from arrest to ED arrival), ED factors (initial rhythm at ED arrival and ED CPR duration), and outcomes (return of spontaneous circulation [ROSC] in the ED, survival to discharge or inter-hospital transfer, and favorable neurologic outcome). A favorable neurologic outcome was defined as survival to discharge with a Cerebral Performance Category (CPC) score of 1 or 2; patients who died were classified as having an unfavorable outcome, whereas survivors without a recorded CPC score were classified as having an unknown outcome.

The population denominators were derived from monthly resident registration data by single year of age, which were obtained from the Korean Statistical Information Service (KOSIS) (16). Incidence is expressed per 100,000 person-years.

### Missing data

Missing data were not imputed. Descriptive analyses were based on the available cases for each variable, and the number of missing or unknown values for each variable is reported in the tables. The incidence analyses, which were based on counts of registry entries, had no missing data because age and ED arrival date were available for all included cases.

## Statistical analysis

Continuous variables are presented as medians (interquartile ranges [IQRs]) and were compared using the Kruskal–Wallis test; categorical variables were compared using the χ² test. Crude incidence is presented with exact Poisson 95% confidence intervals (CIs). For the ITS analysis (17), monthly OHCA counts were modeled using segmented Poisson regression, with the log of the age-specific population as an offset. The models included terms for the underlying time trend, level and slope changes at the start of Phase II and Phase III, and Fourier terms for seasonality. Standard errors were adjusted for overdispersion and autocorrelation (18), and the results are reported as incidence rate ratios (IRRs) with 95% CIs. Differences across age groups were tested using age-group-by-intervention interaction terms. The sensitivity analyses used negative binomial regression, omitted the seasonal terms, used a 6-month lag for the standard error adjustment, shifted both phase boundaries one month earlier or later, started the series in 2009, and restricted the analysis to the resuscitation-attempted cohort. Exploratory analyses were performed by pediatric age subgroup and etiology (disease-related vs injury-related). All tests were two-sided (α = 0.05). Analyses were performed using Python 3.13 (statsmodels 0.15).

### Ethics statement

This study was approved by the Institutional Review Board of Seoul St. Mary’s Hospital (IRB No. 2025-0963-0001), which waived the requirement for informed consent because only de-identified registry data were used.

## Results

### Study population

Of the 456,839 registry entries, 10,965 pediatric OHCA cases were included in the incidence analysis, and 6,891 pediatric patients in whom resuscitation was attempted formed the clinical cohort (Phase I, 5,370; Phase II, 817; Phase III, 704). The crude incidence of pediatric OHCA was 7.04 (95% CI 6.90– 7.19), 7.14 (95% CI 6.75–7.55), and 8.35 (95% CI 7.86–8.87) per 100,000 person-years in Phases I, II, and III, respectively, and the corresponding rates of resuscitation-attempted OHCA were 4.38, 4.63, and 5.48 per 100,000 person-years (Table 1).

**Table 1.** Demographic data of pediatric out-of-hospital cardiac arrest.

|  | **Phase I**<br>**(n = 5,370)** | **Phase II**<br>**(n = 817)** | **Phase III**<br>**(n = 704)** | **P**<br>**value** |
|---|---|---|---|---|
| All EMS-assessed OHCA, n | 8,632 | 1,261 | 1,072 |  |
| All EMS-assessed OHCA, rate<sup>*</sup> | 7.04 (6.90–7.19) | 7.14 (6.75–7.55) | 8.35 (7.86–8.87) |  |
| Resuscitation-attempted OHCA, rate<sup>*</sup> | 4.38 (4.27–4.50) | 4.63 (4.32–4.96) | 5.48 (5.09–5.90) |  |
| Age, years, median (IQR) | 6 (0–15) | 7 (0–15) | 10 (1–15) | 0.011 |
| Age group |  |  |  | <0.001 |
| Infant (<1 yr) | 1,551 (28.9) | 261 (31.9) | 174 (24.7) |  |
| Child (1–11 yr) | 1,819 (33.9) | 214 (26.2) | 228 (32.4) |  |
| Adolescent (12–18 yr) | 2,000 (37.2) | 342 (41.9) | 302 (42.9) |  |
| Sex |  |  |  | <0.001 |
| Male | 3,426 (63.8) | 507 (62.1) | 392 (55.7) |  |
| Female | 1,944 (36.2) | 310 (37.9) | 312 (44.3) |  |
| Any documented comorbidity |  |  |  | <0.001 |
| Yes | 275 (13.3) | 65 (21.9) | 35 (13.4) |  |
| No | 1,792 (86.7) | 232 (78.1) | 227 (86.6) |  |
| Missing/unknown | 3,303 | 520 | 442 |  |

<sup>\*</sup>Incidence per 100,000 person-years (95% CI).
Values are n (%); percentages were calculated among patients with known values, and the numbers of patients with missing or unknown values are shown separately.
OHCA, out-of-hospital cardiac arrest; EMS, emergency medical services; IQR, interquartile range; CI, confidence interval.

In the clinical cohort, the age distribution differed across phases. The proportion of adolescents increased from 37.2% in Phase I to 41.9% in Phase II and 42.9% in Phase III, and the proportion of female patients was higher in Phase III than in Phase I (44.3% vs 36.2%)(Table 1).

### Characteristics of pediatric OHCA

Home residence was the most common location of arrest (61.9%, 68.0%, and 70.1% of cases with a known location in Phases I, II, and III, respectively), whereas the proportion of arrests on streets or highways decreased (14.4%, 10.4%, and 5.5%) and that of arrests in ambulances increased (2.7%, 5.0%, and 7.7%). Among disease-related arrests, cardiac origin was the most common etiology (63.7%, 56.9%, and 64.5%), followed by sudden unexpected infant death (SUID; 28.8%, 33.9%, and 27.5%). Among injury-related arrests, the proportion of falls increased (20.5%, 35.2%, and 43.9%), whereas that of traffic accidents decreased (31.1%, 18.5%, and 11.3%). Suicide accounted for the majority of arrests with an intentional cause in all phases (91.7%, 93.9%, and 95.9%; P = 0.238), and the proportion of all injury-related arrests attributed to suicide increased (19.0%, 26.9%, and 30.4%)(Table 2).

**Table 2.** Characteristics of pediatric out-of-hospital cardiac arrest.

|  | **Phase I<br>(n = 5,370)** | **Phase II<br>(n = 817)** | **Phase III<br>(n = 704)** | **P<br>value** |
|---|---|---|---|---|
| **Location** |  |  |  | <0.001 |
| Home residence | 3,031 (61.9) | 458 (68.0) | 455 (70.1) |  |
| Healthcare facility | 175 (3.6) | 18 (2.7) | 13 (2.0) |  |
| Public/Commercial building | 356 (7.3) | 30 (4.5) | 34 (5.2) |  |
| Street/Highway | 705 (14.4) | 70 (10.4) | 36 (5.5) |  |
| Place of recreation | 77 (1.6) | 5 (0.7) | 7 (1.1) |  |
| Ambulance | 132 (2.7) | 34 (5.0) | 50 (7.7) |  |
| Other | 423 (8.6) | 59 (8.8) | 54 (8.3) |  |
| Missing/unknown, n | 471 | 143 | 55 |  |
| **Etiology** |  |  |  |  |
| **Disease** |  |  |  | <0.001 |
| Cardiac origin | 1,741 (63.7) | 230 (56.9) | 202 (64.5) |  |
| SUID | 787 (28.8) | 137 (33.9) | 86 (27.5) |  |
| Respiratory origin | 44 (1.6) | 21 (5.2) | 11 (3.5) |  |
| Other | 159 (5.8) | 16 (4.0) | 14 (4.5) |  |
| **Injury** |  |  |  | <0.001 |
| Choking | 442 (17.7) | 69 (17.5) | 63 (16.6) |  |
| Drug intoxication | 49 (2.0) | 12 (3.0) | 14 (3.7) |  |
| Drowning | 390 (15.6) | 53 (13.4) | 46 (12.1) |  |
| Burns | 37 (1.5) | 0 (0.0) | 0 (0.0) |  |
| Falls | 511 (20.5) | 139 (35.2) | 167 (43.9) |  |
| Hanging/strangulation | 210 (8.4) | 35 (8.9) | 33 (8.7) |  |
| Traffic accidents | 777 (31.1) | 73 (18.5) | 43 (11.3) |  |
| Injury, other | 80 (3.2) | 14 (3.5) | 14 (3.7) |  |
| Missing/unknown, n | 26 | 7 | 8 |  |
| **Intent** |  |  |  | 0.238 |
| Suicide | 478 (91.7) | 108 (93.9) | 118 (95.9) |  |
| Assault | 43 (8.3) | 7 (6.1) | 5 (4.1) |  |
| Unintentional or undetermined, n | 2,001 | 287 | 265 |  |
| **First responder type** |  |  |  | <0.001 |
| Unknown | 1,573 (29.3) | 175 (21.4) | 95 (13.5) |  |
| EMS | 239 (4.5) | 57 (7.0) | 71 (10.1) |  |
| Healthcare provider | 185 (3.4) | 31 (3.8) | 35 (5.0) |  |
| Lay person | 268 (5.0) | 79 (9.7) | 89 (12.6) |  |
| Acquaintance | 274 (5.1) | 37 (4.5) | 23 (3.3) |  |
| Family | 2,831 (52.7) | 438 (53.6) | 391 (55.5) |  |
| **Bystander CPR** |  |  |  | 0.204 |
| Yes | 841 (77.6) | 281 (80.5) | 237 (74.8) |  |
| No | 243 (22.4) | 68 (19.5) | 80 (25.2) |  |
| Missing/unknown, n | 3,879 | 382 | 281 |  |
| **Initial cardiac rhythm at ED** |  |  |  | <0.001 |
| Shockable | 112 (2.1) | 7 (0.9) | 7 (1.0) |  |
| Non-shockable | 3,678 (68.9) | 679 (84.1) | 594 (84.9) |  |
| Undetermined rhythm | 1,547 (29.0) | 121 (15.0) | 99 (14.1) |  |
| Missing/Unknown, n | 33 | 10 | 4 |  |
| **Time of arrest to arrival at ED, min, median (IQR)** | 25 (18–35) | 31 (24–40) | 27 (19–35) | <0.001 |
| Missing, n | 3,716 | 529 | 398 |  |
| **CPR duration in ED, min, median (IQR)** | 30 (20–42) | 30 (21–42) | 30 (21–39) | 0.445 |
| Missing, n | 92 | 0 | 0 |  |

Values are n (%); percentages were calculated among patients with known values, and the numbers of patients with missing or unknown values are shown separately.
SUID, sudden unexpected infant death; EMS, emergency medical services; CPR, cardiopulmonary resuscitation; ED, emergency department; VF, ventricular fibrillation; VT, ventricular tachycardia; PEA, pulseless electrical activity.

Family members were the most common first responders (52.7%, 53.6%, and 55.5%); the proportions of lay persons (5.0%, 9.7%, and 12.6%) and EMS personnel (4.5%, 7.0%, and 10.1%) increased, whereas the proportion with an unknown first responder type decreased (29.3%, 21.4%, and 13.5%). Bystander CPR status was unknown in most cases; among cases with a known status, the rate of bystander CPR did not differ significantly across phases (P = 0.204). Non-shockable rhythms predominated at ED arrival (68.9%, 84.1%, and 84.9%), and shockable rhythms were rare (2.1%, 0.9%, and 1.0%). The median time from arrest to ED arrival was longer in Phase II than in Phase I (31 vs 25 minutes; P < 0.001), whereas ED CPR duration did not differ across phases; however, the time of arrest was missing for most patients (Table 2).

### Clinical outcomes

In the pediatric clinical cohort, the rate of ROSC in the ED was 37.2%, 31.6%, and 30.4% in Phases I, II, and III, and the rate of survival to discharge or transfer was 16.5%, 10.4%, and 10.4%, respectively (both P < 0.001). Favorable neurologic outcomes were rare and did not differ across phases (1.4%, 0.9%, and 1.3%; P = 0.509) (Table 3).

**Table 3.** Outcomes of pediatric out-of-hospital cardiac arrest.

|  | **Phase I**<br>**(n = 5,370)** | **Phase II**<br>**(n = 817)** | **Phase III**<br>**(n = 704)** | **P**<br>**value** |
|---|---|---|---|---|
| **ROSC in the ED** |  |  |  | <0.001 |
| Yes | 2,000 (37.2) | 258 (31.6) | 214 (30.4) |  |
| No | 3,370 (62.8) | 559 (68.4) | 490 (69.6) |  |
| **Survival to discharge or transfer** |  |  |  | <0.001 |
| Yes | 886 (16.5) | 85 (10.4) | 73 (10.4) |  |
| No | 4,477 (83.5) | 732 (89.6) | 631 (89.6) |  |
| Missing/unknown, n | 7 | 0 | 0 |  |
| **Favorable neurologic outcome (CPC 1–2)** |  |  |  | 0.509 |
| Yes | 64 (1.4) | 7 (0.9) | 9 (1.3) |  |
| No | 4,659 (98.6) | 808 (99.1) | 695 (98.7) |  |
| Missing/unknown, n | 647 | 2 | 0 |  |

Values are n (%); percentages were calculated among patients with known values, and the numbers of patients with missing or unknown values are shown separately.
ROSC, return of spontaneous circulation; CPC, Cerebral Performance Category.

### Interrupted time-series analysis of pediatric OHCA incidence

Figure 1 and Table 4 present the ITS results for pediatric OHCA. Before the pandemic, incidence showed a slight upward trend (annual IRR 1.006 [95% CI 1.001–1.012]; P = 0.032). At the implementation of social distancing in March 2020, incidence decreased immediately by 7.4% (IRR 0.926 [95% CI 0.865–0.991]; P = 0.027), with no significant change in slope (monthly IRR 1.003 [95% CI 1.000–1.007]; P = 0.081). After restrictions were lifted, incidence increased immediately (IRR 1.138 [95% CI 1.048–1.235]; P = 0.002) and subsequently declined gradually (monthly IRR 0.993 [95% CI 0.988–0.999]; P = 0.023).

**Figure 1.**
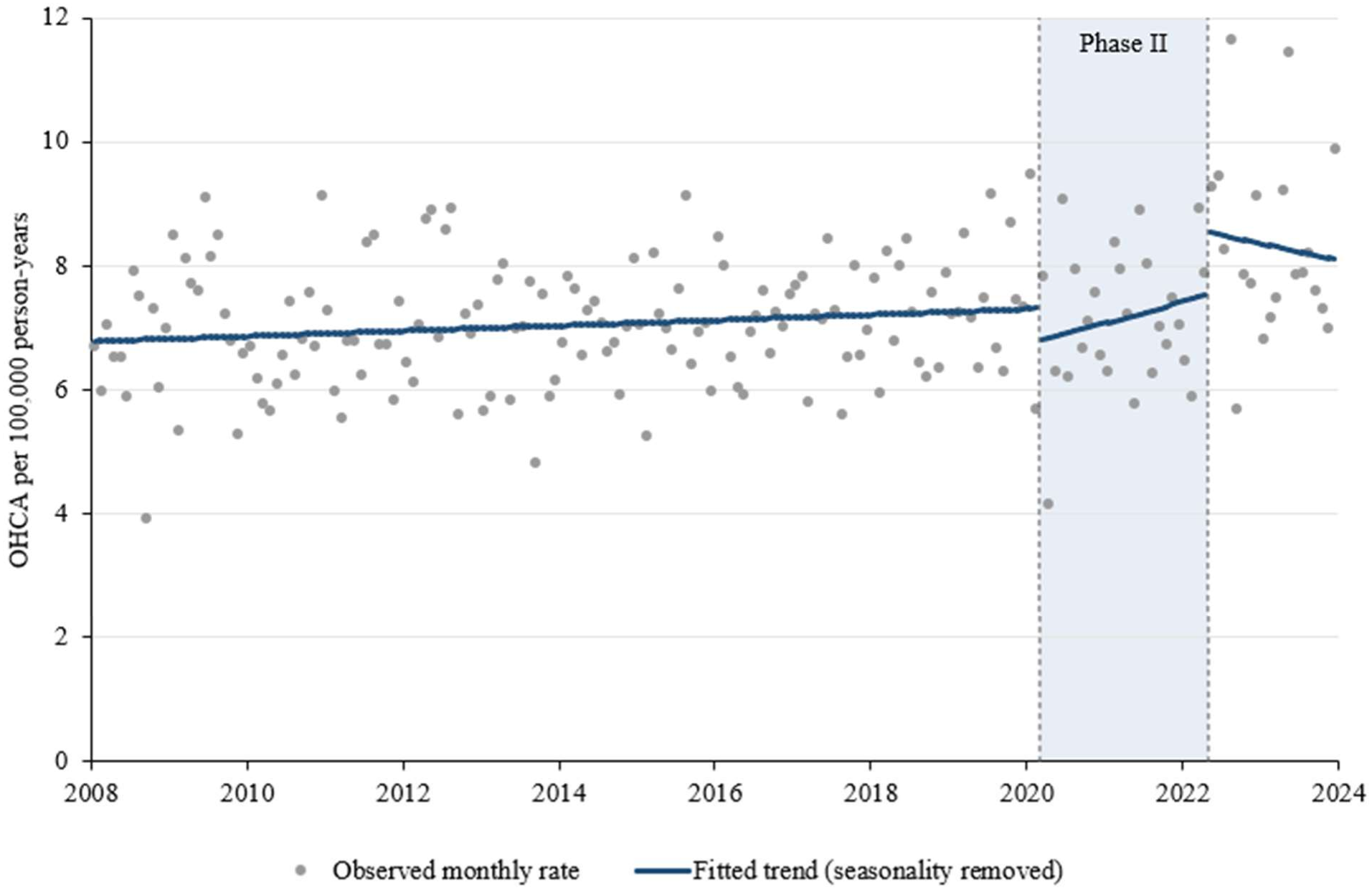
Monthly incidence of pediatric out-of-hospital cardiac arrest in Korea, 2008–2023. Dots show the observed monthly incidence (per 100,000 person-years), and solid lines show the trends fitted by segmented Poisson regression after the removal of seasonality. The shaded area indicates the social distancing period (March 2020–April 2022).

**Table 4.** Interrupted time-series analyses of out-of-hospital cardiac arrest incidence by age group.

|  | **Pediatric<br>IRR (95% CI)** | **P value** | **Adult<br>IRR (95% CI)** | **P value** | **Elderly<br>IRR (95% CI)** | **P value** |
|---|---|---|---|---|---|---|
| Pre-intervention trend (annual IRR) | 1.006 (1.001–1.012) | 0.032 | 1.005 (0.998–1.013) | 0.138 | 1.006 (0.995–1.016) | 0.274 |
| Phase II level change | 0.926 (0.865–0.991) | 0.027 | 0.974 (0.921–1.030) | 0.357 | 0.905 (0.838–0.976) | 0.010 |
| Phase II slope change (monthly IRR) | 1.003 (1.000–1.007) | 0.081 | 1.002 (1.000–1.005) | 0.073 | 1.005 (1.002–1.008) | <0.001 |
| Phase III level change | 1.138 (1.048–1.235) | 0.002 | 0.981 (0.945–1.018) | 0.306 | 0.908 (0.869–0.950) | <0.001 |
| Phase III slope change (monthly IRR) | 0.993 (0.988–0.999) | 0.023 | 0.997 (0.995–1.000) | 0.031 | 0.992 (0.989–0.995) | <0.001 |

Estimates are from segmented Poisson regression of monthly counts of EMS-assessed OHCA with a population offset and seasonal terms; 95% CIs and P values are based on standard errors adjusted for overdispersion and autocorrelation. Level changes are IRRs for the first month of each phase relative to the projected preceding trend; slope changes are monthly IRRs relative to the preceding phase.
IRR, incidence rate ratio; CI, confidence interval; EMS, emergency medical services; OHCA, out-of-hospital cardiac arrest.

In the subgroup analyses (Supplementary Table 1), these changes were confined to children aged 1– 11 years, in whom incidence decreased at implementation (IRR 0.812 [95% CI 0.666–0.991]) and increased after lifting (IRR 1.562 [95% CI 1.278–1.909]). By contrast, no significant level changes were observed in infants (implementation IRR 1.052 [95% CI 0.862–1.284]; post-lifting IRR 0.979 [95% CI 0.808–1.187]) or adolescents (implementation IRR 1.030 [95% CI 0.889–1.195]; post-lifting IRR 1.003 [95% CI 0.882–1.140]). After adjustment for multiple comparisons, the post-lifting increase in children aged 1–11 years remained significant (q < 0.001), whereas the decrease at implementation did not (q = 0.134). By etiology, the post-lifting increase was observed for disease-related arrests (IRR 1.210 [95% CI 1.031–1.420]; P = 0.019) but not for injury-related arrests (IRR 1.032 [95% CI 0.913– 1.165]; P = 0.615).

### Comparison across age groups

The characteristics and outcomes of adult and elderly patients with OHCA are presented in Supplementary Tables 2 and 3, respectively. The policy-associated changes differed significantly across age groups (P < 0.001; Figure 2, Table 4). In adults, no immediate change was observed either at implementation (IRR 0.974 [95% CI 0.921–1.030]; P = 0.357) or after lifting (IRR 0.981 [95% CI 0.945–1.018]; P = 0.306). In elderly individuals, incidence decreased at implementation (IRR 0.905 [95% CI 0.838–0.976]; P = 0.010), the slope increased during Phase II (monthly IRR 1.005 [95% CI 1.002–1.008]; P < 0.001), and incidence decreased again after lifting (IRR 0.908 [95% CI 0.869– 0.950]; P < 0.001), followed by a declining slope (monthly IRR 0.992 [95% CI 0.989–0.995]; P < 0.001).

**Figure 2.**
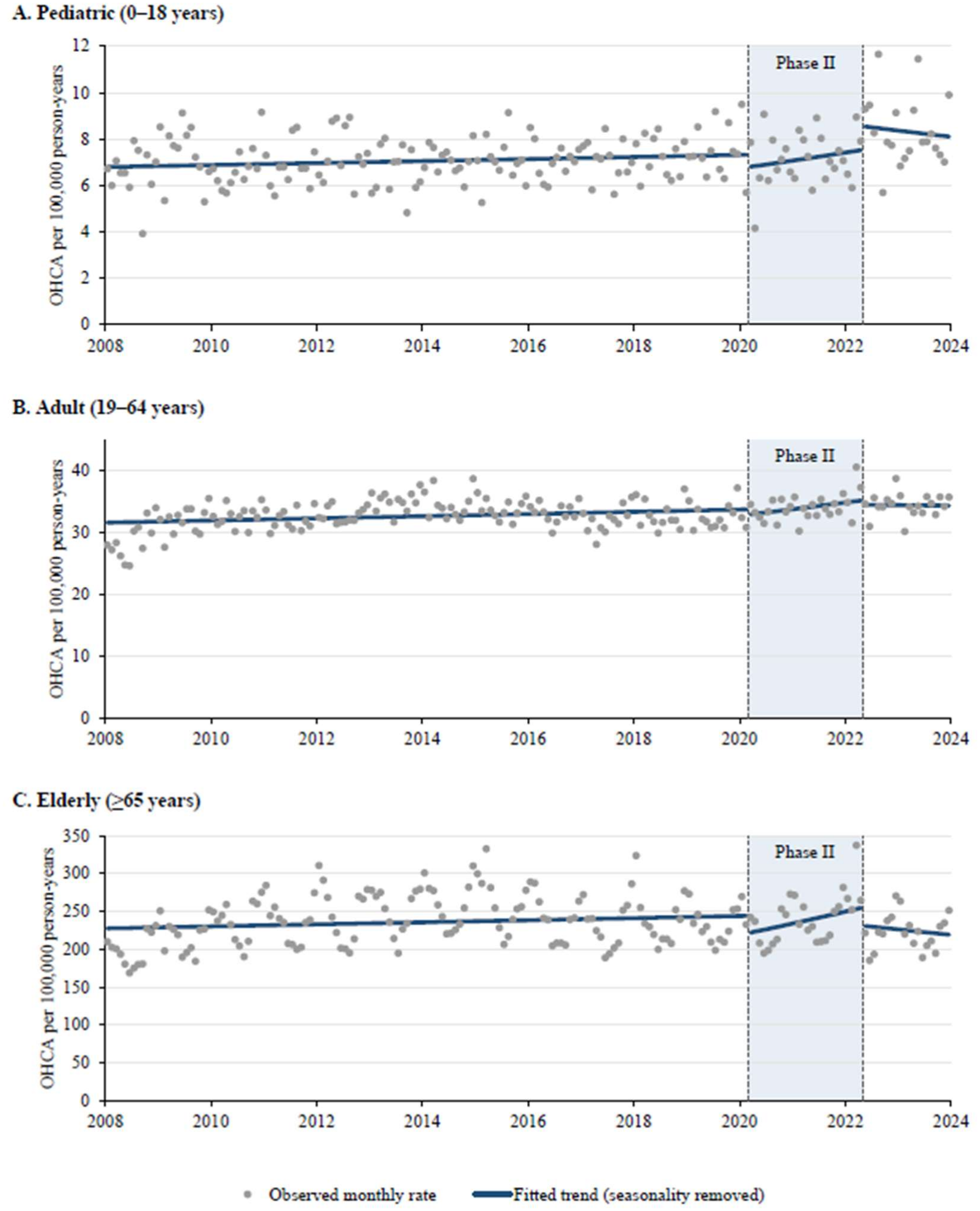
Monthly incidence of out-of-hospital cardiac arrest by age group, 2008–2023. (A) Pediatric (0–18 years), (B) adult (19–64 years), and (C) elderly (≥65 years) populations. Dots show the observed monthly incidence (per 100,000 person-years), and solid lines show the trends fitted by segmented Poisson regression after the removal of seasonality. The shaded area indicates the social distancing period (March 2020–April 2022).

The immediate decrease at implementation did not differ significantly between children and adults (ratio of IRRs 1.052 [95% CI 0.950–1.164]; P = 0.333) or between children and elderly individuals (ratio of IRRs 0.977 [95% CI 0.870–1.096]; P = 0.689). In contrast, the level change after lifting differed markedly across groups: relative to that in children, the post-lifting IRR was lower in both adults (ratio of IRRs 0.862 [95% CI 0.790–0.940]; P < 0.001) and elderly individuals (ratio of IRRs 0.798 [95% CI 0.732–0.871]; P < 0.001).

### Sensitivity analyses

The direction of the pediatric estimates was consistent across all sensitivity analyses, although the Phase II level decrease was of borderline significance in some specifications (P = 0.060–0.074) and the Phase III level increase was not significant when the boundary was shifted one month later (P = 0.188). Differences across age groups remained significant in every scenario (all P < 0.001; Supplementary Table 4).

## Discussion

In this nationwide 16-year study of pediatric OHCA in Korea, three main findings emerged. First, the incidence of EMS-assessed pediatric OHCA was largely stable before the pandemic, increasing by less than 1% per year. Second, the implementation of social distancing was associated with a immediate decrease of approximately 7.4% in pediatric OHCA incidence, which did not persist as a sustained change in trend. Third, after restrictions were lifted, the incidence of pediatric OHCA rebounded above the expected level, and both the decrease and the rebound were concentrated among children aged 1– 11 years. It was the post-lifting rebound, rather than the response at implementation, that distinguished children from the adult and elderly populations.

The near-flat pre-pandemic trend is consistent with previous reports of stable pediatric OHCA incidence in North America (1). Over the same period, the pediatric population in Korea declined substantially, which makes incidence estimates sensitive to the choice of denominator. We therefore used the age-specific monthly resident registration population by single year of age and applied the same approach consistently across all age groups.

The immediate decrease at implementation is biologically and behaviorally plausible. The closure of schools and childcare facilities, reduced outdoor activity and travel, and reduced transmission of respiratory pathogens would be expected to lower exposure to the external and infectious triggers that precede many pediatric arrests (7, 19, 20). The confinement of the decrease to children aged 1–11 years supports this interpretation, because this age group experienced the greatest change in daily routine through the closure of schools and childcare facilities, whereas the circumstances of infant arrests, which largely occur at home and include SUID, would be expected to change little. The absence of a sustained change in slope indicates that the effect was transient, a pattern consistent with the progressive relaxation of school closures and the tiered adjustment of restrictions during the policy period.

The rebound after restrictions were lifted is a novel finding. The incidence of pediatric OHCA increased immediately in May 2022 and declined only gradually thereafter, again driven by children aged 1–11 years. In the exploratory analyses, this increase was observed for disease-related rather than injury-related arrests, although this difference did not remain significant after adjustment for multiple comparisons. This timing coincides with the resumption of in-person schooling and group activities and with the resurgence of common pediatric respiratory infections after a prolonged period of reduced circulation. Although these data cannot establish causality, the pattern suggests that the population-level protection afforded by social distancing may have been offset by a period of increased vulnerability once exposures resumed. Because Phase III spanned only 20 months and its level change was sensitive to shifting the boundary one month later, this finding warrants confirmation with longer follow-up.

The comparison across age groups showed that the immediate decrease at implementation was of similar magnitude in children and elderly individuals but was not detected in adults, and that the difference in policy-associated changes across age groups was driven mainly by the post-lifting period, when pediatric incidence rose while elderly incidence fell. The decrease in elderly OHCA after lifting may reflect changes in healthcare use, a shift toward in-hospital rather than out-of-hospital deaths, or competing mortality during the pandemic; however, this decrease was not the focus of the present study.

Beyond incidence, the etiologic composition of pediatric OHCA shifted across phases. Among injury-related arrests, the proportions attributed to falls and suicide increased, whereas that of traffic accidents decreased; the proportion of adolescents also increased. Suicide accounted for more than 90% of arrests with an intentional cause in all phases. Adolescent suicide is a major public health concern in Korea (21), and the rising contribution of suicide and falls to pediatric OHCA highlights the need for youth mental health and injury prevention strategies. In addition, ROSC and survival to discharge or transfer were lower during and after the social distancing period, and arrest-to-ED intervals were longer in Phase II. These findings are consistent with reported pandemic-related changes in EMS operations and healthcare access (8, 9), although the high proportion of missing prehospital time data limits their interpretation. Favorable neurologic outcomes were rare and did not change across phases.

A key strength of this study is the use of KOHCAR, a nationwide, government-mandated registry that captures all EMS-assessed OHCA cases in Korea, whereas registries such as the European Registry of Cardiac Arrest and the Pan-Asian Resuscitation Outcomes Study vary in participation and data completeness across countries (22, 23). Combined with a clearly defined national policy timeline, this allowed us to estimate changes at both the implementation and the withdrawal of social distancing. Methodologically, the ITS models used population offsets, were adjusted for seasonality, and used standard errors robust to overdispersion and autocorrelation; moreover, the post-lifting rebound was robust across nearly all sensitivity analyses.

This study has several limitations. First, as a registry-based observational study, it cannot establish causality, and other concurrent changes, such as pandemic waves, changes in healthcare use, and the resurgence of respiratory infections after 2022, may have contributed to the observed patterns. Second, the category definitions were derived from registry codes; thus, misclassification of etiology, location, and first responder type is possible. Third, data on several prehospital variables, including bystander CPR and time of arrest, were substantially missing. Finally, Phase III was relatively short (20 months), and the subgroup analyses were exploratory.

In conclusion, the incidence of pediatric OHCA in Korea was largely stable before the COVID-19 pandemic, decreased transiently after the implementation of nationwide social distancing, and rebounded above the expected level after the restrictions were lifted, with these changes concentrated among children aged 1–11 years. Continuous national surveillance that accounts for abrupt changes in children’s daily environments, together with injury prevention and youth mental health strategies, may help reduce the burden of pediatric OHCA.

## Data availability

KOHCAR data are available from the KDCA upon reasonable request and with its approval. The analysis code is available from the corresponding authors upon request.

## CRediT authorship contribution statement

In Young Choi: Data curation, Formal analysis, Visualization, Writing – original draft. Yonghyuk Jeon: Conceptualization, Methodology, Writing – original draft.

In Kyung Lee: Conceptualization, Methodology, Writing – original draft, Writing – review & editing.

Kyunghoon Kim: Conceptualization, Methodology, Resources, Supervision, Writing – review & editing.

All authors had access to the underlying data, contributed to the interpretation of the findings, approved the final version of the manuscript, and agree to be accountable for all aspects of the work.

